# Determinants of Developing Symptomatic Disease in Ethiopian COVID-19 Patients

**DOI:** 10.1101/2020.10.09.20209734

**Authors:** Tigist W. Leulseged, Degu G. Alemahu, Ishmael S. Hassen, Endalkachew H. Maru, Wuletaw C. Zewde, Nigat W. Chamesew, Kalkidan T. Yegile, Daniel S. Abebe, Firaol M. Abdi, Etsegenet Y. Menyelshewa, Tegenu G. Gerbi, Helen T. Hagos

## Abstract

**Background:** Studies show that having some symptoms seems to be associated with more severe disease and poor prognosis. Therefore, knowing who is more susceptible to symptomatic COVID-19 disease is important to provide targeted preventive and management practice. The aim of the study was to assess the determinants of having symptomatic disease among COVID-19 patients admitted to Millennium COVID-19 Care Center in Ethiopia.

**Methods:** A case-control study was conducted from August to September 2020 among a randomly selected 765 COVID-19 patients (372 Asymptomatic and 393 Symptomatic patients). Chi-square test and independent t-test were used to detect the presence of a statistically significant difference in the characteristics of the cases (symptomatic) and controls (asymptomatic), where p-value of <0.05 considered as having a statistically significant difference. Multivariable binary logistic regression was used to assess a statistically significant association between the independent variables and developing symptomatic COVID-19 where Adjusted Odds ratio (AOR), 95% CIs for AOR, and P-values were used for testing significance and interpretation of results.

**Results:** The result of the multivariable binary logistic regression shows that age group (AOR= 1.818, 95% CI= 1.210, 2.731, p-value=0.004 for 30-39 years; AOR= 1.611, 95% CI= 1.016, 2.554, p-value=0.043 for 40-49 years and AOR= 4.076, 95% CI= 2.582, 6.435, p-value=0.0001 for years and above), sex (AOR= 1.672, 95% CI= 1.216, 2.299, p-value=0.002) and history of diabetes mellitus (AOR= 2.406, 95% CI= 1.384, 4.181, p-value=0.002) were found to be significant factors that determine the development of symptomatic disease in COVID-19 patients.

**Conclusions:** Developing a symptomatic COVID-19 disease was found to be determined by exposures of old age, male sex, and being diabetic. Therefore, patients with the above factors should be given enough attention in the prevention and management process, including inpatient management, to pick symptoms earlier and to manage accordingly so that these patients can have a favorable treatment outcome.

## INTRODUCTION

The new coronavirus disease 2019 (COVID-19) caused by a virus in the coronaviruses family called severe acute respiratory syndrome coronavirus 2 (SARS-CoV-2) was first identified in China in December 2019 and declared to be a pandemic by the World Health Organization in March 2020^1^.

Till today, there is no such pathognomonic feature identified for the disease presentation. But according to studies, the COVID-19 disease could present with different symptoms and signs. Regarding the spectrum of symptoms, studies show that the presentation spectrum could range from having no symptoms at all while being infected (asymptomatic cases) to different types of one or more symptoms. Respiratory and constitutional symptoms are reported to be the commonest presentation in most setups^2-4^.

Also, atypical presentations like hyposmia, hypogeusia, nasal congestion, rhinorrhea, sputum, cerebrovascular accidents, abdominal pain, vomiting, and diarrhea are reported. These atypical symptoms are reported to be observed in the majority of patients in a study conducted in Europe but only in few patients in the United States making it difficult to generalize these symptoms as atypical for all setups ^2-8^.

Studies about what determines becoming symptomatic are lacking but different studies and reports point that acquiring the disease, developing symptomatic infection, having severe disease and bad prognosis shows a difference in terms of sex showing that the male sex is more vulnerable to worse conditions. Different explanations are given starting from the genetic level up to behavioral factors that could cause men to be more susceptible to the disease ^9-15^.

Studies including the ones conducted in our center showed that having some symptoms seems to be associated with more severe disease and poor prognosis rather than other symptoms or have no symptoms at all. Therefore, knowing who is more susceptible to symptomatic COVID-19 disease is important to provide a better preventive practice and stratified patient management and follow up. But there is no such study conducted in the study area.

Therefore, the objective of this study was to assess the determinants of having symptomatic disease among COVID-19 patients admitted to Millennium COVID-19 Care Center in Ethiopia.

## METHODS AND MATERIALS

### Study setting and period

The study was conducted at Millennium COVID-19 Care Center (MCCC), a makeshift hospital in Addis Ababa, Ethiopia.

### Study Design

Institution based case-control study design was employed.

### Source and Study Population

The source population was all patients admitted to MCCC with a confirmed diagnosis of COVID-19 using RT-PCR by a laboratory given mandate to test such patients by the Ethiopian Federal Ministry of Health^16^.

– **Case (symptomatic):** all RT-PCR confirmed COVID-19 patients admitted to the MCCC and who presented with one or more symptoms at admission and during the clinical course of the disease.
– **Control (asymptomatic):** all RT-PCR confirmed COVID-19 patients admitted to the MCCC and who did not have any symptoms during the clinical course of the disease.

The study population was all selected COVID-19 patients who were on treatment and follow up at MCCC and who full fill the inclusion criteria.

### Sample Size Determination and Sampling Technique

The sample size was determined using sample size calculation for a case-control study with the following assumptions; 95% confidence interval, power of 90%, one to one ratio of cases and controls, the proportion of males who are symptomatic as 0.85, and proportion of females who are asymptomatic as 0.75 and considering a non-response rate of 10%. Therefore, the total sample size calculated becomes 778 cases of COVID-19 (389 symptomatic and 389 asymptomatic).

To select both the cases and the controls, simple random sampling method was used.

### Eligibility criteria

All COVID-19 patients who were on treatment and follow up at MCCC and whose clinical history regarding presenting symptom and symptoms developed during the clinical course of the disease is well documented were included.

### Operational Definition

#### Asymptomatic patient

any patient who has tested positive for COVID-19 but does not have any symptoms. These patients are detected after isolation and contact tracing as done by Ethiopian Public Health Institute ^16^.

### Data Collection Procedures and Quality Assurance

A data abstraction tool to pick all the relevant variables was drafted based on the patient registration and follow-up form and then pretested on 5% of randomly selected charts which were not included in the final data collection. The final data abstraction tool was then developed after modification based on the pretest. Data was collected by six trained General practitioners.

Data consistency and completeness were checked before an attempt was made to enter the code and analyze the data.

### Data Management and Analysis

The extracted data were coded, entered into Epi-Info version 7.2.1.0, cleaned, stored, and exported to SPSS version 25.0 software for analysis. Categorical covariates were summarized using frequencies and percentages and numerical variables were summarized with a mean value. A chi-square test (Fischer’s exact test for those variables that do not meet the chi-square test assumptions) and an independent t-test were run to determine the presence of a significant difference between the independent variables and the developmentof symptom. The assumptions of the chi-square test, that no cell should have an expected frequency of less than 5, and the assumptions of t-test, normal distribution, and equality of variance, were checked before the analysis, and all assumptions were met. A statistically significant difference was detected for variables with a P-value of ≤ 0.05.

The association between the dependent variable and independent variables was analyzed using multivariable Binary Logistic Regression. Univariate analysis was done at a 25% level of significance to screen out independent variables used in the multiple Binary Logistic regression model. The adequacy of the final model was assessed using the Hosmer and Lemeshow goodness of fit test and the final model fitted the data well (x^2^(7)=6.298 and p-value = 0.505). For the Binary Logistic regression, a 95% confidence interval for AOR was calculated and variables with p-value ≤ 0.05 were considered as statistically associated with the development of symptoms (Symptomatic Vs Asymptomatic).

## RESULT

### Socio–demographic, co-morbid illness and disease related variables and comparison between Symptomatic Vs Asymptomatic

From the 778 charts, information was collected from 765 charts making the response rate 98.3%. The majorities (25.9%) of the patients involved in the study were in the two extreme age groups, < 30 years (33.9%) and ≥ 50 years (26.3%). Four hundred fifty (56.8%) were males. Close to one third (29.8%) of the patients had a history of one or more pre-existing co-morbid illness. The majority had hypertension (16.6%), diabetes mellitus (11.9%), cardiac disease (3.7%), and Asthma (3.4%). One hundred forty-four (18.8%) patients needed oxygen therapy.

Based on the chi-square/ Fischer’s exact test result, a significant difference in the presence of symptom was found among the different age groups, sex, different COVID-19 severity, those with a history of pre-existing co-morbid illness, cardiac disease, hypertension, diabetes mellitus, asthma and the need of oxygen supplement.

Accordingly, a significantly higher proportion of patients in the age group of < 30 years were asymptomatic (66.5 % Vs 33.5%, p-value=0.0001). On the other hand for the rest of the three age groups, a significantly higher proportion of patients were symptomatic. The majority of females were asymptomatic (55.9% Vs 44.1%, p-value=0.001) and males were symptomatic (43.6% Vs 56.4%, p-value=0.001). A greater proportion of moderate (83.3% Vs 16.7%, p-value=0.0001) and severe (96.8% Vs 24.4%, p-value=0.0001) patients were symptomatic and the mild patients were asymptomatic (83.4% Vs 16.6%, p-value=0.0001). The majority of patients with pre-existing co-morbid illness (71.1% Vs 28.9%, p-value=0.0001) were symptomatic. Similarly, patients with cardiac disease, hypertension, diabetes, and asthma were symptomatic compared with those patients with no such illness. Those who needed oxygen therapy (97.2% Vs 2.8%, p-value=0.0001) were symptomatic compared to those who was on room air. (**Table 1**)

**Table 1:**
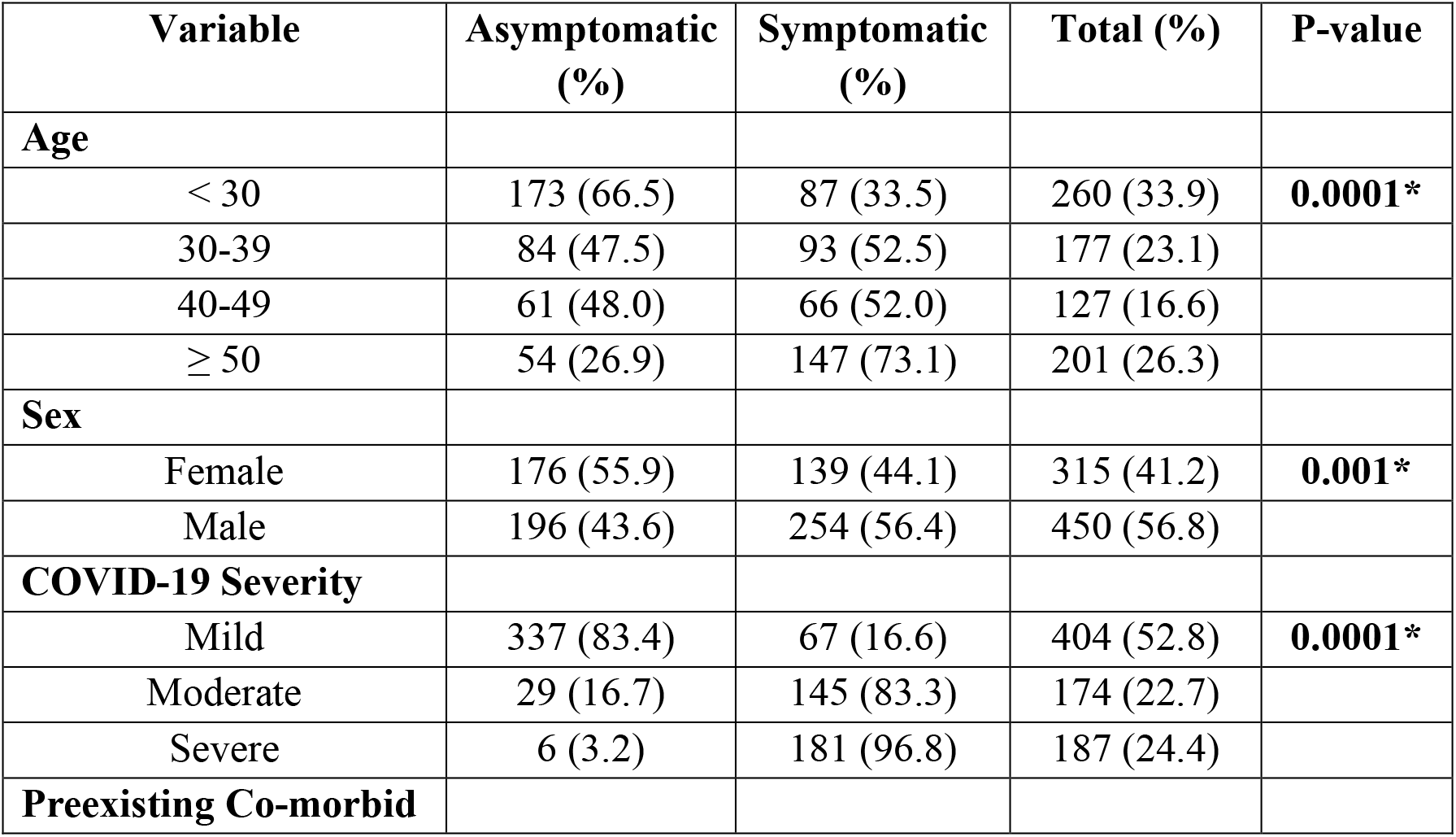
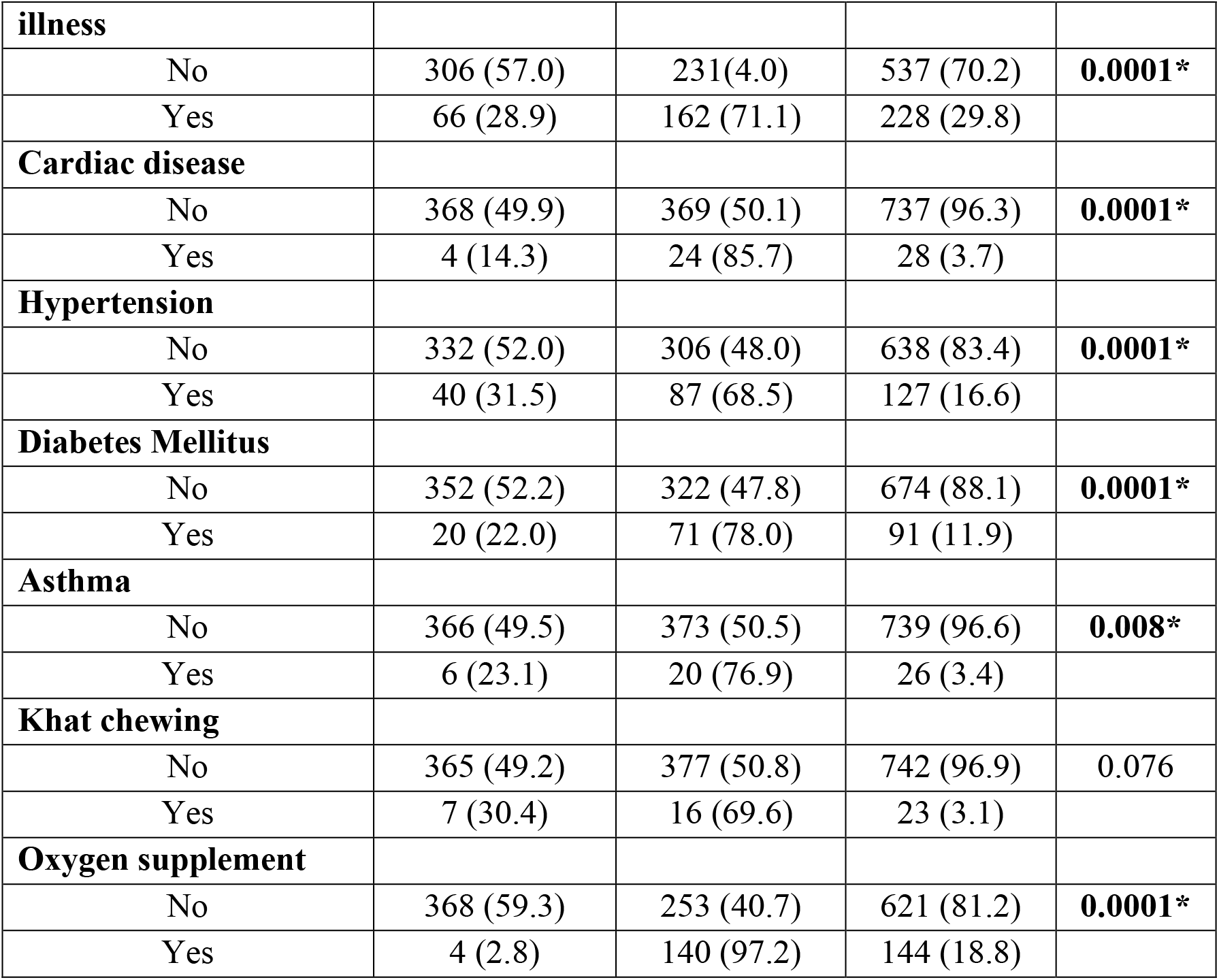
Socio–demographic, preexisting co-morbid illness and disease related variables and comparison between Symptomatic Vs Asymptomatic COVID-19 patients (n=765)

### Baseline vital sign and laboratory markers related variables and comparison between Symptomatic Vs Asymptomatic

Based on the results of the independent t-test, symptomatic patients were relatively tachypnic (19.9/min Vs 23.2/min, p-value=0.0001) and have lower oxygen saturation (95.6% Vs 93.7%, p-value=0.0001). In addition, the symptomatic patients had a neutrophil predominant cell count (78.3% Vs 57.7%, p-value=0.005)

Symptomatic patients have a significantly raised urea (22.9 mg/dl Vs 35.7 mg/dl, p-value=0.027) and alanine transaminase (21.6 IU/L Vs 49.9 IU/L, p-value=0.005). On the other raised aspartate transaminase was seen in the asymptomatic patients (53.9 IU/L Vs 44.9%, p-value=0.007). (**Table 2**)

**Table 2:** Baseline vital sign and Laboratory markers related variables and comparison between Symptomatic Vs Asymptomatic COVID-19 patients (n=765)

| Variable | Asymptomatic | Symptomatic | P-value |
| --- | --- | --- | --- |

|  | (Mean) | (Mean) |  |
| --- | --- | --- | --- |
| **Vital sign** |  |  |  |
| Temperature (°C) | 37.5 | 37.2 | 0.786 |
| Heart rate (Beats/min) | 97.1 | 97.3 | 0.446 |
| Respiratory rate (RR/min) | 19.9 | 23.2 | **0.0001\*** |
| SBP (mmHg) | 135.8 | 129.9 | 0.116 |
| DBP (mmHg) | 83.6 | 79.6 | 0.231 |
| Spo2 (%) | 95.6 | 93.7 | **0.0001\*** |
| **Complete blood count** |  |  |  |
| Hemoglobin (mg/dl) | 14.9 | 15.0 | 0.568 |
| Hematocrit (%) | 43.6 | 43.9 | 0.635 |
| White blood cell count (Cells/L) | 7.9 | 26.2 | 0.443 |
| Neutrophil % (%) | 57.7 | 78.3 | **0.005\*** |
| Absolute neutrophil count<br>(cells/L) | 540.7 | 740.3 | 0.436 |
| Lymphocyte % (%) | 21.7 | 12.0 | 0.550 |
| Platelet count (cells/ L) | 363.1 | 267.8 | 0.268 |
| Hemoglobin (mg/dl) | 315.56 | 271.8 | 0.313 |
| **Renal function test** |  |  |  |
| Urea (mg/dl) | 22.9 | 35.7 | **0.027\*** |
| Creatinine (mg/dl) | 1.12 | 1.21 | 0.901 |
| **Liver function test** |  |  |  |
| Asparatate transaminase (IU/L) | 53.9 | 44.9 | **0.007\*** |
| Alanine transaminase (IU/L) | 21.6 | 49.4 | **0.005\*** |
| Alkaline phosphatase (IU/L) | 87.4 | 89.0 | 0.261 |
| **Electrolyte** |  |  |  |
| Sodium (mEq/L) | 138.8 | 142.7 | 0.528 |
| Potassium (mEq/L) | 4.4 | 4.3 | 0.081 |

### Factors associated with the presence of symptom in COVID-19 patients

Univariate analysis at 25% level of significance was conducted and age group, sex, hypertension, and diabetes mellitus history were found to be significantly associated with the development of symptoms.

On the multivariable binary logistic regression, age group, sex, and diabetes mellitus were found to be significantly associated with the development of symptoms at 5% level of significance.

Accordingly, after adjusting for other covariates, the odds of developing symptomatic disease was more likely among patients in the age range of 30 years and above compared with those patients < 30 years (AOR= 1.784, 95% CI= 1.189, 2.675, p-value=0.005 for 30-39 years; AOR= 1.650, 95% CI= 1.043, 2.609, p-value=0.032 for 40-49 years and AOR= 4.196, 95% CI= 2.666, 6.604, p-value=0.005 for years and above).

The odds of developing symptomatic COVID-19 among males was 1.674 times than female patients (AOR= 1.674, 95% CI= 1.219, 2.298, p-value=0.001).

Regarding the history of pre-existing co-morbid illness, the odds of developing symptomatic disease among diabetic patients was 2.368 times compared to patients with no such illness (AOR= 2.368, 95% CI= 1.364, 4.111, p-value=0.002). (**Table 3**)

**Table 3:**
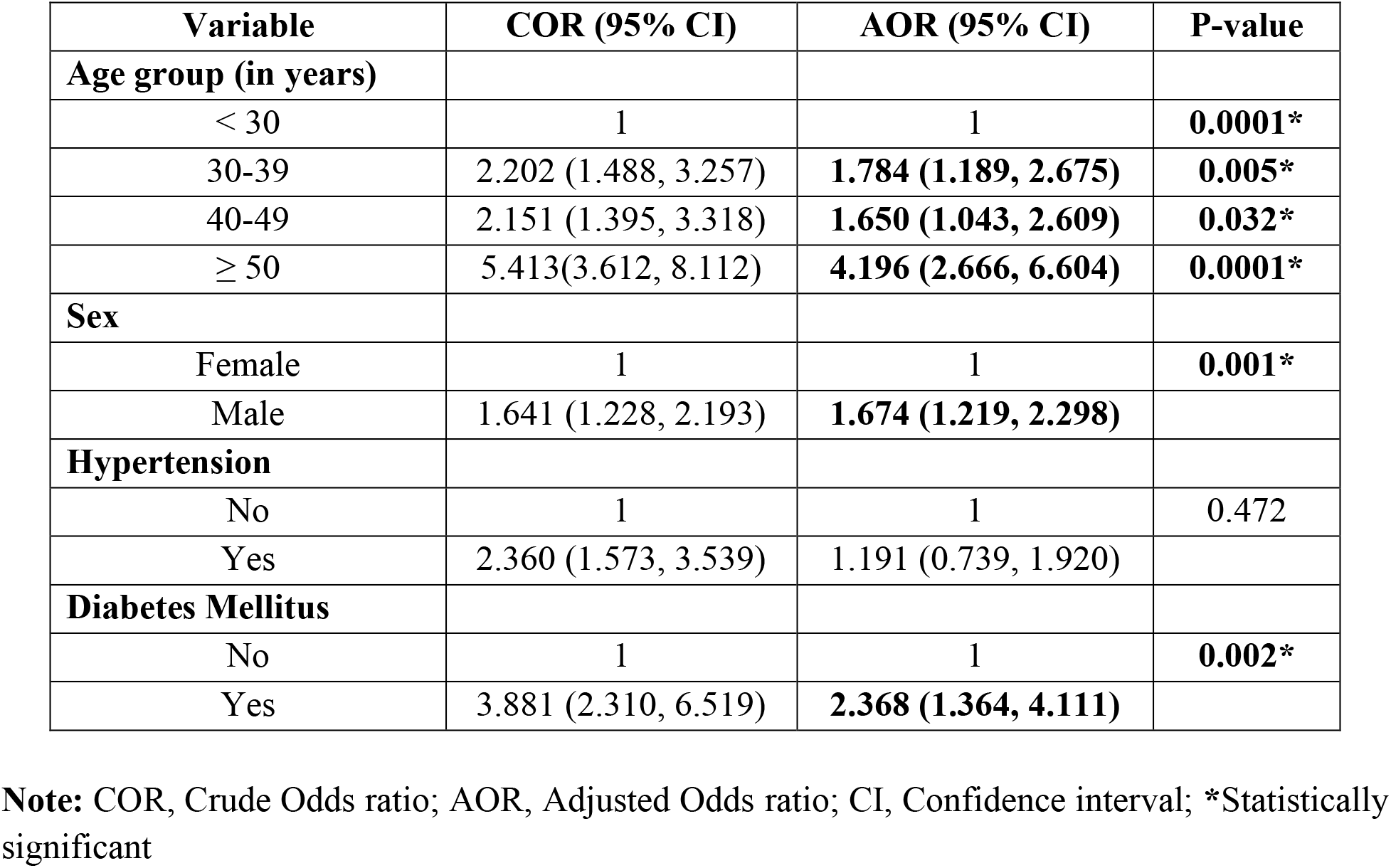
Results for the final multivariable binary logistic regression model among COVID-19 patients (n=765)

## DISCUSSION

This study has tried to assess the determinants of developing symptomatic COVID-19 disease among RT-PCR confirmed patients admitted at MCCC. The chi-square test result of the study shows that a significant difference in having symptomatic Vs asymptomatic infection is observed in some of the variables showing that symptomatic disease is significantly associated with old age, male sex, having moderate and severe disease, having one or more co-morbid illness and requiring oxygen therapy. Based on the t-test, symptomatic infection is significantly associated with a relatively higher respiratory rate, lower oxygen saturation, neutrophil predominance, raised urea and alanine transaminase, and lower aspartate transaminase.

On the multivariable binary logistic regression; age group, sex, and diabetes mellitus were significantly associated with the development of symptoms.

Older age was found to be a significant factor that affects the development of symptomatic COVID-19. Accordingly, after adjusting for other covariates, the odds of developing symptoms were more likely among patients in the age range of 30 years and above compared with those patients < 30 years old. The odds are higher (more than four times) for those patients 50 years and above. That means older age groups are more susceptible to developing symptomatic disease once they are infected. This could be because, as it is already known, the body’s immune defense mechanism tends to get weaker with age making aged people more susceptible to severe disease and worse outcomes from both infectious and non-infectious diseases. This is especially compounded by the presence of other underlying conditions (comorbidities) which are highly likely to be developed as someone gets older. The participants included in the study have also a significantly higher proportion of patients with comorbid illnesses that can be attributed to the increased odds of developing symptomatic disease^17-20^.

Male sex was associated with symptomatic COVID-19 infection. The odd of having symptomatic COVID-19 among males was 1.672 times than female patients. This disparity in sex in terms of symptoms and also acquiring the disease, having a more severe disease course, and worse outcome is observed in many countries with reports showing that a significantly higher proportion of men are getting hospitalized and dying from the disease compared to females. Different explanations have been given to the disparity, from genetics up to behavioral factors as contributors. Studies show that Angiotensin-converting enzyme 2 (ACE2) which is found to be a receptor for the SARS-CoV-2 is found to be present in high concentration among males, which makes them vulnerable to have a more severe disease pattern and worse outcome ^9-15^. It is also hypothesized that men might not be as good as women in terms of taking necessary precautions to prevent acquiring the disease and also in seeking early medical attention if they get infected and become symptomatic. Understanding these modifiable behavioral factors is important to provide intervention to decrease severe disease and worse outcomes in men.

The other important factor that determines the development of symptomatic infection was being a diabetic patient. The odds of developing symptomatic disease among diabetic patients were 2.368 times compared to patients with no such illness. It is well known that diabetes mellitus is one of the chronic medical illnesses that is associated with depressed body immunity that will leave the patient to be susceptible to any infectious diseases. It also increases the patients’ probability of developing another comorbid illness which in turn could result in diabetes disease progression and development of short and long-term complications including poor glycaemic control that will make the patient more susceptible to external invaders resulting in symptomatic infection, severe disease, and poor outcome. This is also found to be the case in other studies, where having diabetes is associated with unfavorable clinical presentation and COVID-19 outcome^17-20^.

## CONCLUSION

Developing a symptomatic COVID-19 infection was found to be determined by the following factors; old age, male sex, and being diabetic.

Therefore, patients with the above factors should be given enough attention, including inpatient management, to pick symptoms earlier and to manage accordingly so that these patients can have a favorable treatment outcome. In addition, further study should be conducted to assess the role of behavioral factors as a cause of the disease course disparity between the two sexes, so that intervention can be made on the identified modifiable behaviors.

## Data Availability

All relevant data are available upon reasonable request

## Declaration

### Ethics approval and consent to participate

The study was conducted after obtaining ethical clearance from St. Paul’s Hospital Millennium Medical College Institutional Review Board. Written informed consent was obtained from the participants. The study had no risk/negative consequence on those who participated in the study. Medical record numbers were used for data collection and personal identifiers were not used in the research report. Access to the collected information was limited to the principal investigator and confidentiality was maintained throughout the project.

### Authors Contribution

All authors contributed to the conception of the study. TWL designed the study, revised data extraction sheet, performed statistical analysis, and drafted the initial manuscript. All authors obtained patient data. All authors revised the manuscript and approved the final version.

## Acknowledgment

The authors would like to thank St. Paul’s Hospital Millennium Medical College for facilitating the research work.

## Availability of data and materials

All relevant data are available upon reasonable request.

